# Estimating the increase in reproduction number associated with the Delta variant using local area dynamics in England

**DOI:** 10.1101/2021.11.30.21267056

**Authors:** Sam Abbott, CMMID COVID-19 Working Group, Adam J. Kucharski, Sebastian Funk

## Abstract

**Background:** Local estimates of the time-varying effective reproduction number (*R*_*t*_) of COVID-19 in England became increasingly heterogeneous during April and May 2021. This may have been attributable to the spread of the Delta SARS-CoV-2 variant. This paper documents real-time analysis that aimed to investigate the association between changes in the proportion of positive cases that were S-gene positive, an indicator of the Delta variant against a background of the previously predominant Alpha variant, and the estimated time-varying *R*_*t*_ at the level of upper-tier local authorities (UTLA).

**Method:** We explored the relationship between the proportion of samples that were S-gene positive and the *R*_*t*_ of test-positive cases over time from the 23 February 2021 to the 25 May 2021. Effective reproduction numbers were estimated using the `EpiNow2` R package independently for each local authority using two different estimates of the generation time. We then fit a range of regression models to estimate a multiplicative relationship between S-gene positivity and weekly mean *R*_*t*_ estimate.

**Results:** We found evidence of an association between increased mean *R*_*t*_ estimates and the proportion of S-gene positives across all models evaluated with the magnitude of the effect increasing as model flexibility was decreased. Models that adjusted for either national level or NHS region level time-varying residuals were found to fit the data better, suggesting potential unexplained confounding.

**Conclusions:** Our results indicated that even after adjusting for time-varying residuals between NHS regions, S-gene positivity was associated with an increase in the effective reproduction number of COVID-19. These findings were robust across a range of models and generation time assumptions, though the specific effect size was variable depending on the assumptions used. The lower bound of the estimated effect indicated that the reproduction number of Delta was above 1 in almost all local authorities throughout the period of investigation.

## Introduction

In April 2021, imported cases with the Delta (B.1.617.2) SARS-CoV-2 variant were first detected in England. At the same time, there was increasing concern that the Delta variant had been responsible for the large increase in reported COVID-19 cases in India. In early May, Delta was declared a variant of concern and evidence was required to assess potential differences to the dominant Alpha variant in order to guide policy decisions in England.

To provide evidence rapidly, we adapted previous work on the Alpha variant^[1]^ to estimate the association between the time-varying effective reproduction number (*R*_*t*_) at the level of upper-tier local authorities (UTLA), as estimated using our previously published approach^[2–4]^, and the proportion of positive cases that were S-gene positive. Due to the number of unknowns, we presented a range of models and scenarios exploring model assumptions. Versions of this work were considered on the 2nd and 9th of June by the Scientific Pandemic Influenza Group on Modelling (SPI-M) and by Scientific Advisory Group for Emergencies 91 (SAGE) on the 3rd of June^[5]^. The approach and results presented here have not been substantially updated in order to reflect the evidence available at the time. All data and code used in this study are available with the intention of allowing others to inspect and improve the analysis methodology.

## Method

### Data

We used 4 main sources of data: test positive COVID-19 notifications by UTLA^[6]^, S-gene status from PCR tests by local authority provided by Public Health England (PHE), Google mobility data stratified by context^[7]^, and data on the timings changes in national restrictions^[8]^. National restrictions were coded by hand as either national lockdown or one of the 3 reopening steps used by the UK government during the study period. National lockdown applied prior to the 8th of March 2021 when schools reopened in England (step 1). On the 12th of April outdoor pubs, restaurants and non-essential shops reopened in England (step 2) with this being followed by most rules affecting outdoor social contact being removed, six people being allowed to meet indoors, and indoor hospitality services being reopened on the 17th of May 2021 (step 3).

We aggregated mobility data by taking the mean for each week. We calculated the weekly proportion of positive tests that were S-gene positive over time by local authority by sequence data. We then assumed a one week delay between infection and sequencing date and used this to reference the proportion of S-gene positives by week of infection rather than by week of sequence. Complete case analysis was used and we restricted the data used in later analyses to only include the period from beginning Tuesday, 23 February and ending Tuesday, 25 May. We further restricted the analysis to UTLA/week combinations where more than 20% of reported cases and more than 20 in total had an S-gene status reported.

### Statistical analysis

We estimated reproduction numbers by date of infection using the method described in^[2]^ and^[3]^ and implemented in the `EpiNow2` R package^[4]^. Up to date and archived versions of these estimates can be downloaded at https://github.com/epiforecasts/covid-rt-estimates/blob/master/subnational/united-kingdom-local/cases/summary/rt.csv. We used two sets of estimates, obtained using gamma-distributed generation interval distributions with a mean of 3.6 days (standard deviation (SD): 0.7), and SD of 3.1 days (SD: 0.8)^[2,3,9]^ or with a mean of 5.5 days (SD: 0.5 days), and SD of 2.1 days (SD: 0.25 days)^[10]^, respectively. Weekly estimates of the time-varying reproduction number were then estimated by taking the mean of each daily expected reproduction number in each UTLA for each week. These estimates were then linked to the proportion of cases that were S-gene positive by week of infection and to local restrictions and mobility indicators.

We then built a separate model of the expected reproduction number in UTLA *i* during week *t* starting in the week beginning 23 February, 2021, as a function of local restrictions, mobility indicators, residual temporal variation, and proportion of positive tests S-gene positive:

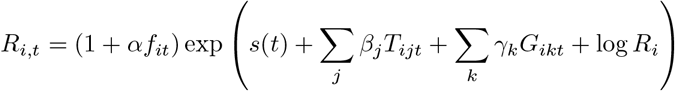

where *R*_*t*_ is an UTLA-level intercept corresponding to *R*_*t*_ during national lockdown in February/March, *T*_*ijt*_ is 1 if intervention *j* (out of: January lockdown, reopening phase 1/2) is in place and 0 otherwise, *G*_*ikt*_ is the relative mobility in context *k* (home, workplace, public transport) at time *t* in UTLA *i* as measured by Google, and *s*(*t*) is a time-varying component, modelled either as a region-specific thin-plate regression spline (“Regional time-varying”), the sum of a static regional parameter and a national spline (“National time-varying”), or only a static regional parameter (“Regional static”). **W**e considered the model with only a static regional parameter as our baseline model as it yielded the most directly interpretable parameter estimates, assuming reproduction numbers could be completely explained by the relaxation steps and spread of S-gene positivity. The spline versions were designed to capture confounding due to unmeasured covariates over time and were therefore considered as lower bounds on effect estimates. We lastly fitted the model for each *t* separately, i.e. at fixed time slices (“Time-sliced”), with and without a regional intercept.

The key parameter is *α*, the relative change in reproduction number in the presence of s-gene positivity that is not explained by any of the other variables, where *f*_*it*_ is the proportion out of all positive tests for SARS-CoV-2 where the S-gene was tested positive, and the reproduction number in any given UTLA is

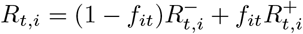

where 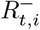 is the S-gene negative reproduction number, 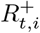 is the S-gene positive reproduction number, and it is assumed that 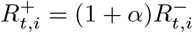.

We used a Student’s t-distribution observation model with a single variance parameter and a single degree of freedom parameter.

### Reproducibility

All models were implemented using the brms^[11]^ package in `R` version 4.0.5^[12]^. Notification data was downloaded and preprocessed using the `covidregionaldata` R package version 0.9.2^[13]^. All code and data required to reproduce this analysis are available from both GitHub and the Open Science Framework.

## Results

Using data from the 23 February 2021 to the 25 May 2021 we found consistent evidence of an association between S-gene positivity and increased UTLA level expected reproduction number estimates. The association became more apparent over time. The outer 90% credible intervals of the estimates in the time-sliced analysis (Fig. 2) ranged from a -10% increase to a 113% increase, in April/May, depending on the assumption about generation times and the model used, as the proportion of tests that were S-gene positive increased heterogeneously across NHS regions. Out of the models fitted to all time, ones that adjusted for residual variation over time on both a national and NHS region level fit the data better than those that did not (Tab. 1). However, all models had evidence of increased *R*_*t*_ with S-gene positivity with the best fitting model yielding a lower bound of 20% higher *R*_*t*_ of S-gene positive cases with a short generation time (Tab. 2), higher than the model that only adjusted for national level residual variation over time (lower bound: 21%) and lower than the model that did not adjust for residual variation over time (lower bound: 28%). With a longer generation time, these lower bounds changed to 27%, 28%, and 38%, respectively. The upper bound of the increase in *R*_*t*_ varied from 33% to 55% in models with different assumed generation times.

**Table 1:** Model comparison (long generation interval) by difference in expected log-predictive density

| Model | ELPD difference | Standard error |
| --- | --- | --- |
| **Short generation time** |  |  |
| Regional time-varying | 0 | 0 |
| National time-varying | -30 | 11 |
| Regional static | -119 | 14 |
| **Long generation time** |  |  |
| Regional time-varying | 0 | 0 |
| National time-varying | -32 | 12 |
| Regional static | -122 | 15 |

**Table 2:** Parameter α with 95% credible intervals for the three different models of s(t) for short (3.6 days mean) and long (5.5 days mean) generation intervals. The estimate corresponds to the multiplicative increase in reproduction number estimated for S-gene positive cases.

| Model | Estimate |
| --- | --- |
| **Short generation time** |  |
| Regional static | 0.28–0.4 |
| National time-varying | 0.21–0.35 |
| Regional time-varying | 0.2–0.33 |
| **Long generation time** |  |
| Regional static | 0.38–0.55 |
| National time-varying | 0.28–0.47 |
| Regional time-varying | 0.27–0.46 |

**Figure 1:**
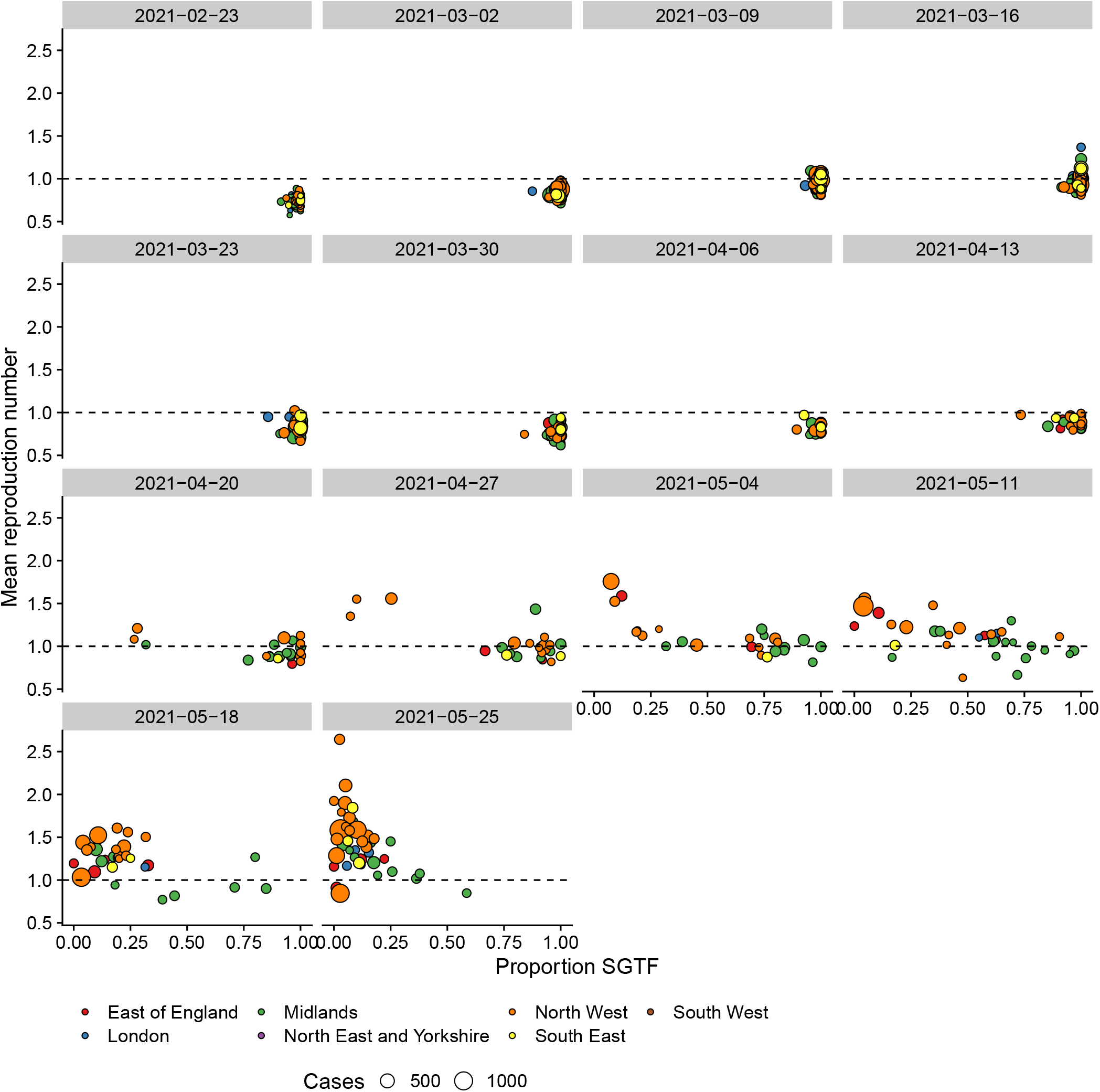
Mean reproduction numbers using a generation time with a mean of 5.5 days since the week beginning 23 February, 2021, compared to the proportion of all test-positives tested for S-gene that tested S-gene positive/negative that week. Each point represents one UTLA, with the size given by the number of cases in the week following the week of the given reproduction number to account for the delay from infection to testing. Only UTLAs with sufficient coverage of S-gene results at least 20% of cases tested and at least 20 results in total)

**Figure 2:**
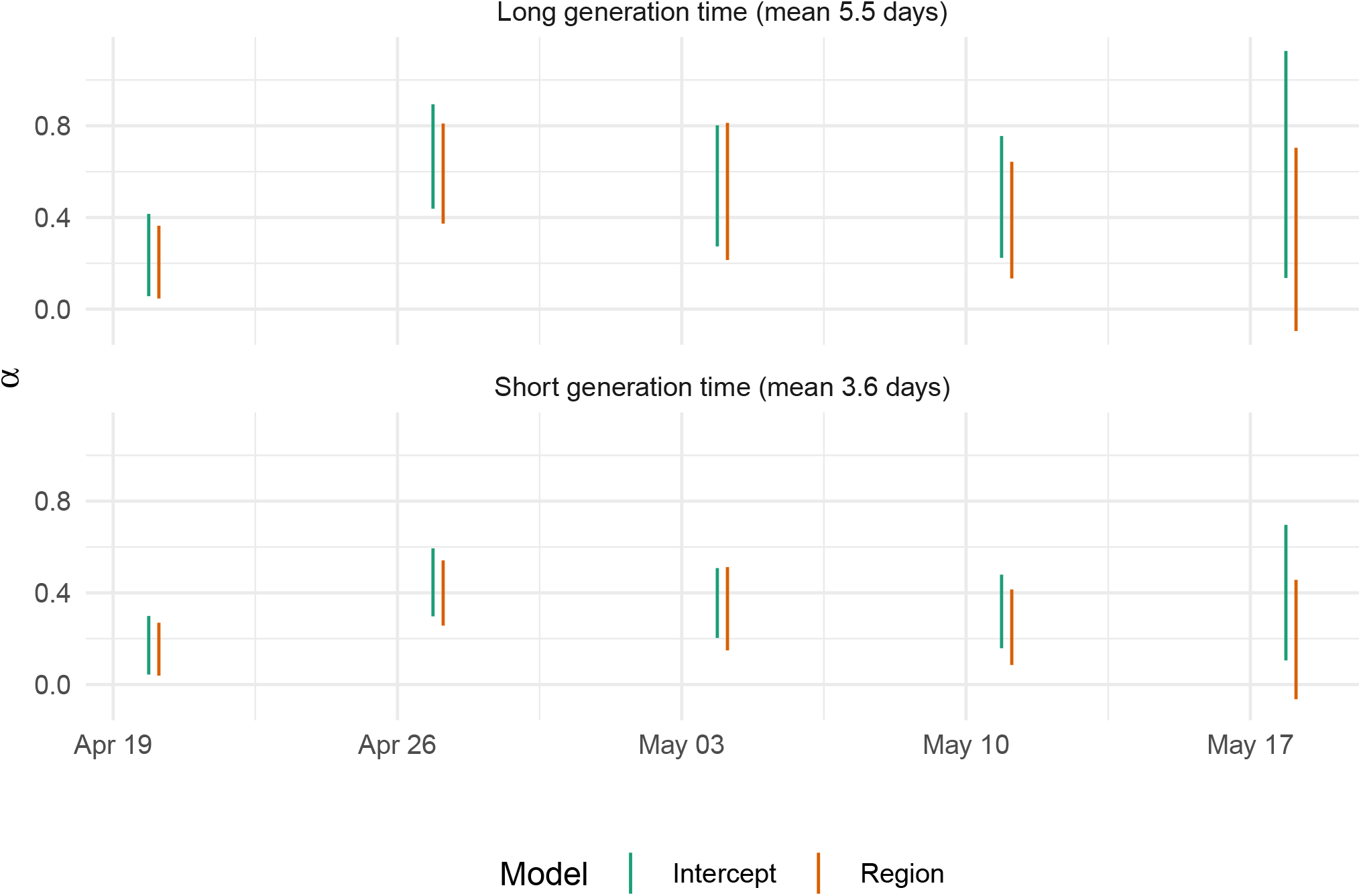
Independent weekly estimates of the multiplicative increase in reproduction number (α) estimated for S-gene positive cases after adjusting for other confounders using the time-sliced model with a global or regional intercept and both a long and a short generation time. 90% credible intervals shown for all estimates.

The model that did not adjust for residual variation appeared to reproduce estimated reproduction numbers relatively well over time (Fig. 3) although there are notable outliers especially in early March, when some of the *R*_*t*_ estimates may have been affected by the results of mass testing in schools entering the case data, and in some of the later weeks when Delta was increasingly prevalent. This model also yielded estimates of the relative impact of the different steps of reopening on *R*_*t*_, with a relatively small effect in the order of 5-20% for each step, but combinedly possibly lifting *R*_*t*_ close to 1 at the time of the third step of reopening even with the previously circulating Alpha variant (Fig. 4).

**Figure 3:**
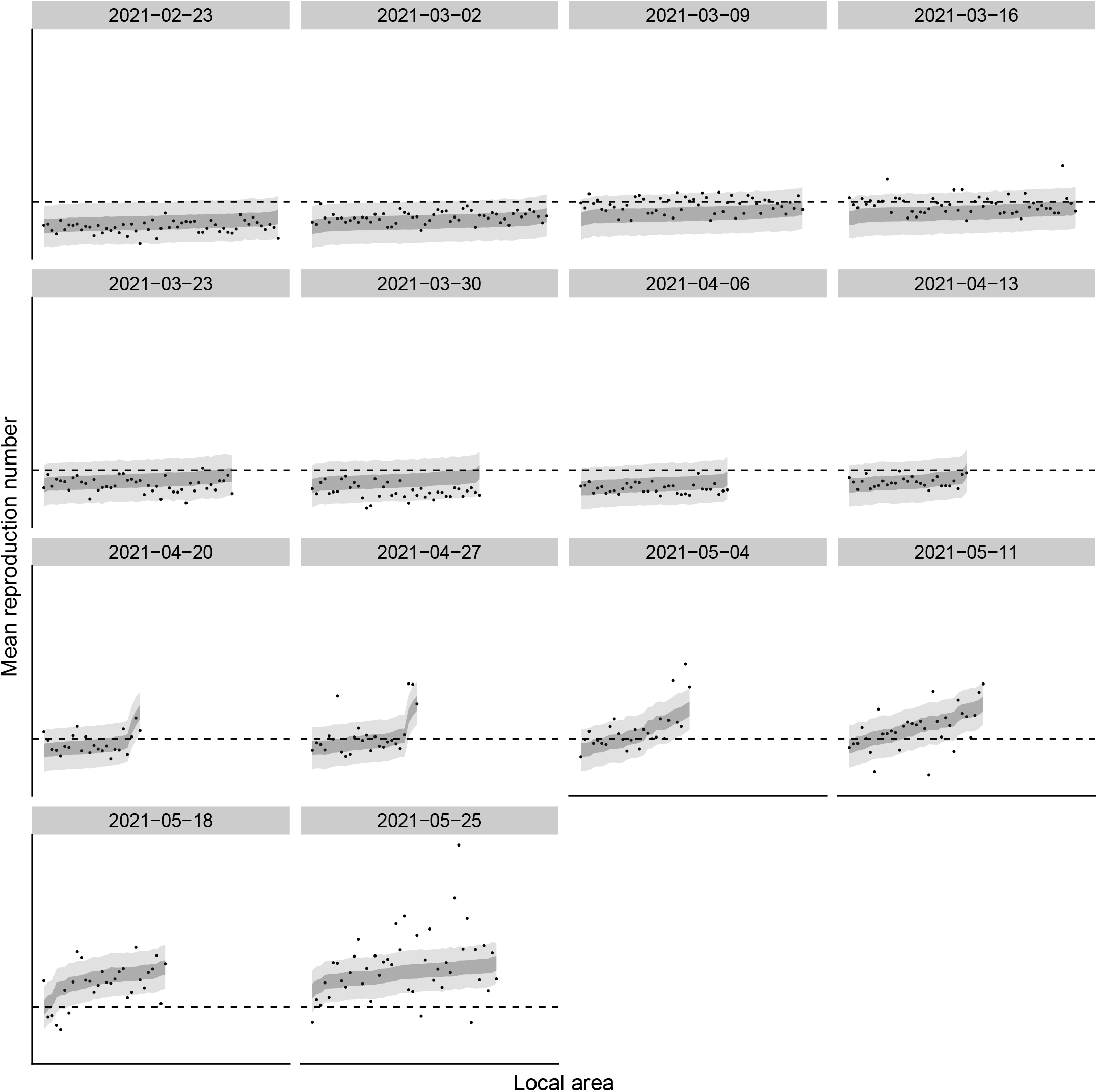
Predictions of the regional static model with long generation interval compared to the data (solid dots). Dark grey: central 50% prediction interval; light grey: central 90% prediction interval. Areas are ordered each week according to predicted median.

**Figure 4:**
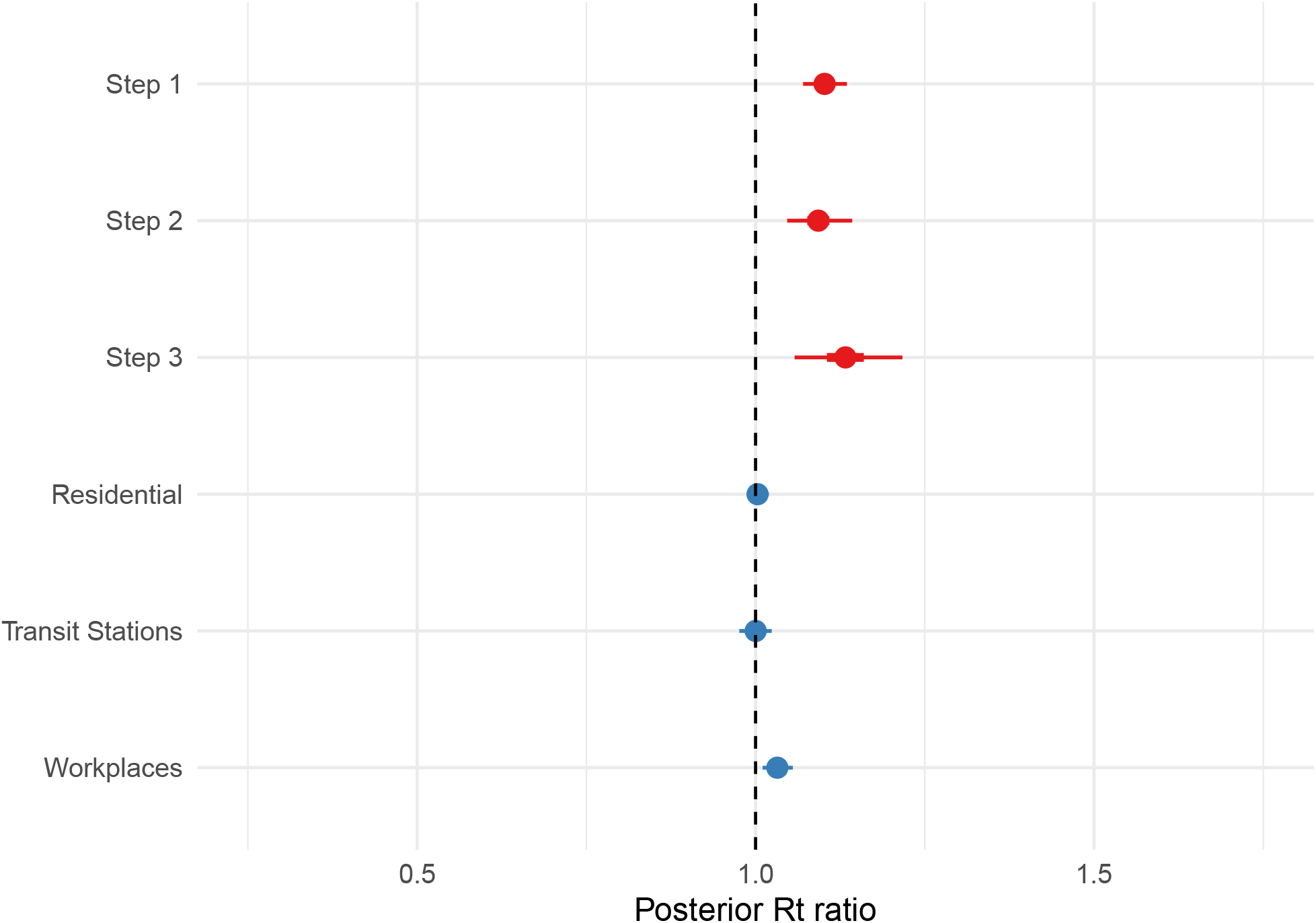
Parameter estimates (R ratios) of the Regional static model. These correspond to a multiplicative effect on a baseline R_t_ of 0.74–0.81. Intervention indicators are coloured in red, and mobility indicators in blue.

## Discussion

We studied the relationship between S-gene positivity (as a proxy for the new variant of concern Delta) and UTLA-level effective reproduction number estimates using four related models that had varying degrees of flexibility in ascribing changes in the effective reproduction number to factors not explained by the proportion of cases with SGTF. The model that associated all temporal variation with interventions, mobility and SGTF produced the largest increases (central estimates: 46% and 34% increase with long and short generation interval, respectively). A more conservative, if more difficult to interpret, model with region-specific splines yielded central estimates of a 26% and 36% increase in *R*_*t*_ with short and long generation interval, respectively. While this model yielded the best fit to the data the estimate of increased *R*_*t*_ associated with SGTF may well be an underestimate as it can explain regional differences by unmodelled factors, i.e. ones beyond interventions, mobility, and distribution of SGTF, and is therefore largely a model of within-region UTLA-level differences. On the other hand, the fact that this model yielded a better fit than the model without splines suggests that that model is affected by confounders not included in the model.

Our estimates that did not adjust for residual variation over time and used the longer generation interval estimate were consistent with ones from other preliminary rapid analyses using similar generation intervals (between 5-6 days), which were in the order of 13-57%^[14]^, 20-60%^[15]^ increase, yet lower than another estimate of 50-100%^[16]^. They are also consistent with increased secondary attack rates from contact tracing during relevant time periods, increased from 8.2% (8.0%-8.4%) for “Alpha” to 12.4% (11.7%-13.2%), corresponding with an increased reproduction number of around 50%^[17]^. That said, other model variants yield lower estimates consistent with the data, as do shorter generation intervals, which generally lead to reproduction numbers closer to 1 and thus lower estimates of a multiplicative effect. This may be particularly relevant where the effect of the variant causes the effective reproduction number to cross 1. In comparing our estimates to others, it is worth bearing in mind the difference between estimating differences in transmission between variants at the national or regional level as many of the other preliminary studies did, vs. the local level as we did. Considering the national picture implicitly gives greater weight to local areas where most cases occur, thus potentially introducing a bias if there are specific conditions that affect spread in these areas, whereas our approach implies weighing all local areas equally in trying to estimate differences in growth by SGTF, thus potentially being more affected by stochastic effects due to small case counts. As it is not clear a priori which of these approaches would yield more accurate estimates of true biological differences, it is useful to consider them in parallel in order to learn from any similarities or differences observed.

Our results should be treated with caution as several caveats apply: we assumed that S-gene positive and negative cases had the same generation interval, while a complementary hypothesis might be that the new variant shortened the generation interval, affecting our estimates^[18]^. We assumed that the effect of tiers and lockdown applied uniformly across the country. While we did allow for a flexible regional-level behaviour through our use of UTLA level intercepts and region-specific regression splines as sensitivity analysis, there may be UTLA level, potentially spatial structured, variation that we did not capture in doing so. If this could explain some of the sub-regional differences in reproduction numbers, our estimate for the increased reproduction number could be biased. In addition, we did not include case importation between UTLAs or cases linked to international travel which may have particularly biased initial estimates. Our estimates are also likely to be overly precise as we fitted the model only to the mean estimated reproduction numbers and therefore ignored uncertainty in these estimates as well as in the proportion of S-gene positives observed in every UTLA per week, which were treated as fixed-point estimates and naively referenced to the week of infection using an assumed delay of one week. Improving our inference method to incorporate these uncertainties is a future aim of our research^[19]^. Our estimates may also be biased as S-gene status is only a proxy for variant identification and the previously predominant Alpha variant may sometimes yield S-gene positives. We therefore cannot rule out that our effect includes a component not related to the new variant. Lastly, our analysis has focussed on the potential for a transmission advantage for the Delta variant but we did not consider alternative mechanisms such as an improved ability to evade immune response elicited by either prior infection or vaccination.

This analysis was done rapidly when Delta started increasing in England, in response to a need to gather scientific evidence for the scale of the transmission advantage and as such represents a real-time rather than a retrospective estimate. The underlying approach used was similar to that used in our previous real-time work on the Alpha’ variant^[1]^ which was also conducted under similar time pressures and had similar methodological limitations. We have not substantially updated the methods or results from the report considered by Scientific Pandemic Influenza Group on Modelling, SPI-M^[5]^ in order to highlight the limitations of our work produced under time pressure and in the absence of data retrospectively available. Future work is ongoing exploring more appropriately modelling uncertainty in both transmission and sequence data and better capturing spatial variation^[19]^. This work should ideally be done with the aim of producing a generalisable approach that can be applied to future scenarios in which rapid evidence needs to be synthesised in order to guide policymakers.

We found consistent evidence that S-gene positivity was associated with increased reproduction numbers across a range of models and assumptions. The precise estimate of the effect size was impacted by both the degree of flexibility allowed in the model used and the assumed generation time. However, the lower bound of the effect implies that the levels of restrictions on contacts present in England through April to June were not sufficient to reduce the reproduction number of the emerging Delta variant to below 1. Our analysis is fully reproducible and all the aggregated data used is publicly available for reuse and reinterpretation.

## Data Availability

All underlying data is available from the Open Science Framework along with code and model output: http://doi.org/10.17605/OSF.IO/H6E8N
Underlying data and code is also available from Zenodo: http://doi.org/10.5281/zenodo.5236662 License: MIT
Source code is available from: https://github.com/epiforecasts/covid19.sgene.utla.rt
Archived source code at the time of publication:
OSF: http://doi.org/10.17605/OSF.IO/H6E8N
Zenodo: http://doi.org/10.5281/zenodo.5236662

https://github.com/epiforecasts/covid19.sgene.utla.rt/tree/v2.0

http://doi.org/10.17605/OSF.IO/H6E8N

http://doi.org/10.5281/zenodo.5236662

## Data availability

This project contains the following underlying raw and processed data:

- `data/utla_rt_with_covariates.rds`: UTLA level weekly reproduction number estimates combined with estimates of the proportion of tests that were S-gene negative/positive, normalised Google mobility data, and tier status by local authority over time.
- `data/rt_weekly.rds`: Summarised weekly UTLA reproduction number estimates using both a short and a long generation time.
- `data/utla_cases.rds`: UTLA level COVID-19 test positive cases.
- `data/sgene_by_utla.rds`: Weekly test positivity data for the S-gene by UTLA.
- `data/mobility.rds`: Normalised Google mobility data stratified by context.
- `data/tiers.rds`: UTLA level tier level over time.

All underlying data is available from the Open Science Framework along with code and model output: http://doi.org/10.17605/OSF.IO/H6E8N

Underlying data and code is also available from Zenodo: http://doi.org/10.5281/zenodo.5236662

License: MIT

## Software availability

Source code is available from: https://github.com/epiforecasts/covid19.sgene.utla.rt Archived source code at the time of publication:

- OSF: http://doi.org/10.17605/OSF.IO/H6E8N
- Zenodo: http://doi.org/10.5281/zenodo.5236662

License: MIT

## Contributors

SA and SF conceived and designed the work, undertook the analysis, and wrote the manuscript. All authors contributed to subsequent drafts. All authors approve the work for publication and agree to be accountable for the work.

## Acknowledgements

The following authors were part of the CMMID COVID-19 Working Group: Simon R Procter, Rachael Pung, Mihaly Koltai, Stéphane Hué, Carl A B Pearson, Katharine Sherratt, Paul Mee, Rosanna C Barnard, Nicholas G. Davies, David Hodgson, Kerry LM Wong, Kaja Abbas, Nikos I Bosse, Yang Liu, Sophie R Meakin, Joel Hellewell, Hamish P Gibbs, Matthew Quaife, Ciara V McCarthy, Yalda Jafari, Frank G Sandmann, Rachel Lowe, James D Munday, Graham Medley, Gwenan M Knight, C Julian Villabona-Arenas, William Waites, Fiona Yueqian Sun, Mark Jit, Alicia Rosello, Akira Endo, Emilie Finch, Christopher I Jarvis, Kiesha Prem, Katherine E. Atkins, Damien C Tully, Lloyd A C Chapman, Oliver Brady, Kathleen O’Reilly, Rosalind M Eggo, and Amy Gimma.

